# Potency and Breadth of Neutralization after 3 doses of mRNA vaccines in COVID-19 Convalescent and Naïve individuals

**DOI:** 10.1101/2022.03.17.22272557

**Authors:** Joanna Luczkowiak, Gonzalo Rivas, Nuria Labiod, Fátima Lasala, Marta Rolo, Jaime Lora-Tamayo, Mikel Mancheno-Losa, David Rial-Crestelo, Alfredo Pérez-Rivilla, María D. Folgueira, Rafael Delgado

## Abstract

Third doses of mRNA COVID-19 vaccines induced a significant increase in neutralizing potency and breadth in naïve individuals comparable with convalescents who restored levels after the first two doses. These results suggest a limit to elicit neutralization in the number of stimuli by infection or vaccination with ancestral SARS-CoV-2 sequences

## Background

The implementation of COVID-19 vaccines has been one of the main tools for pandemic control. The contraction of humoral response after vaccination and the emergence of the different SARS-COV-2 variants of concern (VoC), along with a clear disproportion in the global availability of vaccines, have represented an important challenge to achieve high levels of protection(1). Vaccine coverage has been particularly challenged after the rapid spread of the Omicron VoC which combines an unprecedented number of mutations in its sequence clearly related with increased transmission efficiency and neutralizing scape(2). After the first prime-boost administration of mRNA vaccines, a third dose has shown an important boost of potency and breadth of neutralizing response(3). Here we have investigated the neutralizing response after the third mRNA vaccine dose against SARS-CoV-2 VoC and SARS-CoV-1 in a cohort of COVID-19 convalescent patients in comparison with only-vaccinated naïve individuals.

## Methods

### Participants

We have included 21 COVID-19 convalescent and 21 naïve healthcare workers (HCW) from the Hospital Universitario 12 de Octubre in Madrid, Spain. Both groups were part of a follow-up study (Solidarity II cohort, IRB approval ref CEIm 20/157) and were recruited after informed consent and randomly selected among those with serum samples available for the study period. Mean age was 49 and 48 years for the convalescent and naïve groups, respectively. All infections in convalescent individuals took place during the epidemic wave of COVID-19 affecting Madrid during March-April 2020, and all had a mild clinical evolution. All participants were vaccinated in January-February 2021 with two doses of the Pfizer-BNT162b2 vaccine 21 days apart. Blood samples were obtained at 61 days (range 42-77) and 242 days (range 238-252) after the first dose in the COVID-19 convalescent group and at 67 days (range 49-97) and 241 (range 228-252) in the COVID-19 naïve group. A third dose of 50 ug of the Moderna mRNA-1273 vaccine was administered in December 2021 according with EMA recommendations(4). Participants with documented SARS-CoV-2 infections by RT-PCR or serology (anti-N) were excluded. From the original group, 15 samples from COVID-19 convalescent and 12 from the naïve group were collected 48 days after the third dose (range 41-57).

Production of SARS-CoV-2 pseudotyped VSV: SARS-CoV-2 pseudotyped VSV were produced according to a previously published protocol(5). The SARS-CoV-2 Spike mutant D614G was generated by site-directed mutagenesis using as an input DNA the expression vector encoding SARS-CoV-2 Spike_614D protein (kindly provided by J. Garcia-Arriaza, CNB-CSIC). Spike sequences from SARS-CoV-2 variant Alpha (B.1.1.7, GISAID: EPI_ISL_608430), SARS-CoV-2 variant Beta (B.1.351, GISAID: EPI_ISL_712096), SARS-CoV-2 variant Gamma (P.1, GISAID: EPI_ISL_833140), SARS-CoV-2 variant Delta (B.1.617.2, GISAID: EPI_ISL_1970335) and SARS-CoV-2 variant Omicron (B.1.1.529, GISAID: EPI_ISL_6640917) were synthesized and cloned into pcDNA3.1 by GeneArt technology (Thermo Fisher Scientific GENEART GmbH, Regensburg, Germany). SARS-CoV-1 Spike was purchased from SinoBiological, China (Cat. No. VG40150-G-N).

### Neutralization assay

The neutralization activity of sera was tested as described previously(5). As a control, the neutralization assay included dilution series of an external calibrant with assigned unitage of 813 International Units per ml [IU/ml] for neutralization activity (WHO SARS-CoV-2 Serology International Standard, 20/136, Frederick National Laboratory for Cancer Research)(6).

### RBD-binding antibodies

Anti-RBD Ig titers were determined by an electrochemiluminescence commercial assay (Elecsys Anti-SARS-CoV-2, Roche Diagnostics, Basel, Switzerland) and were converted to WHO International Standard Binding Antibody Units and expressed as BAU/mL following the manufacturer instructions

## Results

Results are described in Figure 1. In COVID-19 naïve individuals, neutralizing titers after prime-boost vaccination with BNT-162b2 were very low against the Omicron VoC (GMT 37 and 31 IU/mL at 2- and 8-months post vaccination respectively) as compared with the group of COVID-19 convalescent individuals (Geometric mean titer, GMT, 802 and 237 IU/mL) (p<0.001). However, the third dose of mRNA 1273 increased the levels of neutralization against Omicron specially in the naïve group that achieved a high and comparable neutralizing potency as the convalescent group (832 vs 894 IU/mL respectively). Neutralizing levels for the same time points for SARS-CoV-2 VoC Alpha, Beta, Gamma and Delta followed a similar pattern as Omicron, significantly lower in naïve individuals after the first two doses (p<0.001) and comparable to convalescents after the third dose. This pattern of response had a correlation with results obtained with RBD-binding total antibodies at the same time points in both groups of participants (Figure 2). The neutralizing titer against SARS-CoV-1 was as expected low for naïve individuals after the first two doses and also increased significantly after the third dose (GMT 397 IU/mL) but this level was significantly lower than the achieved in the convalescent group (GMT 768 IU/mL) (p=0.018). (Figure 1).

**Figure 1.**
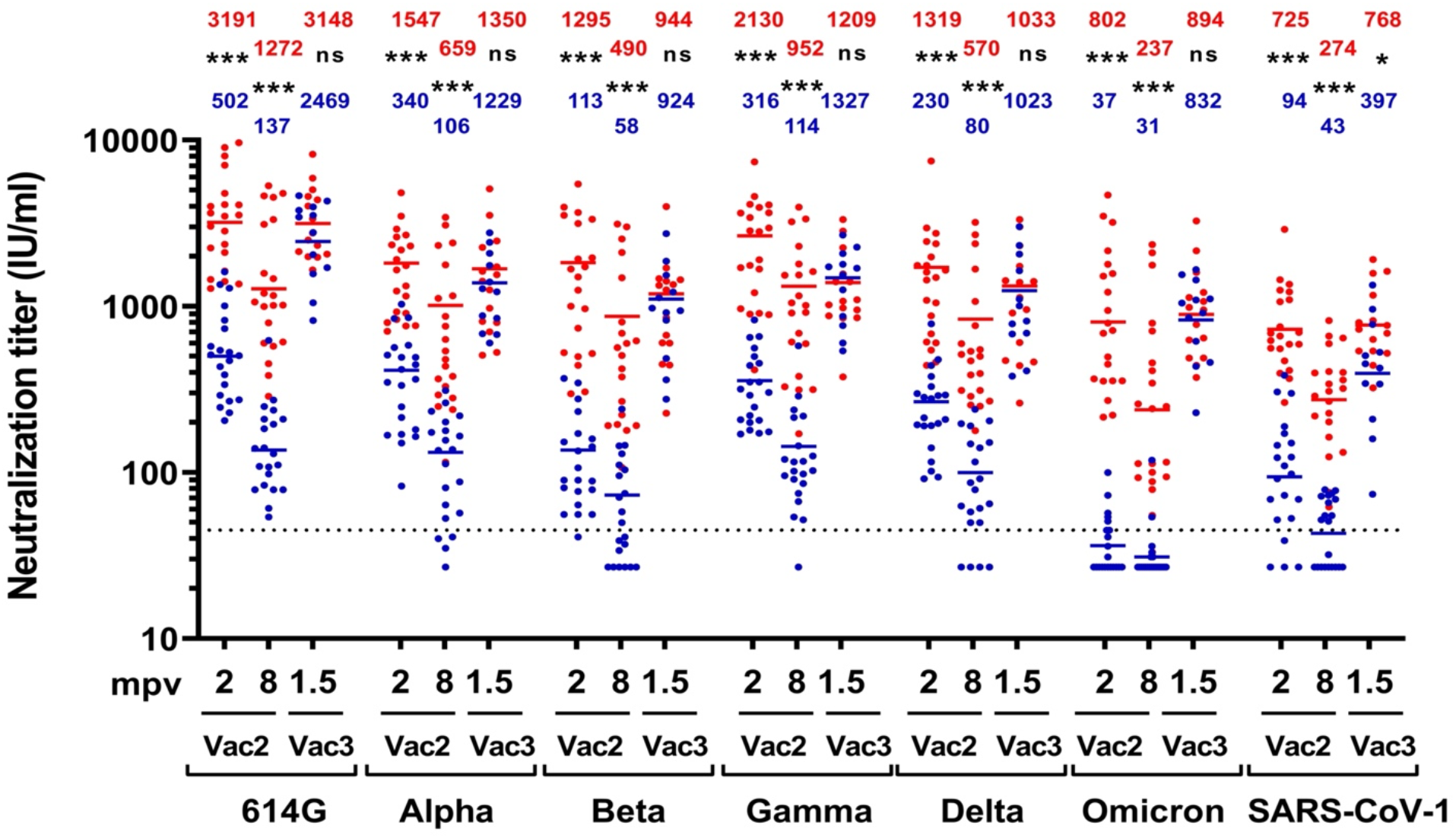
Neutralizing activity against SARS-CoV-2 VoC and SARS-CoV-1 in vaccinated COVID-19 convalescent and naïve individuals. COVID-19 convalescent vaccinated (n=21) individuals are presented as scatter red dot plots and COVID-19 naïve vaccinated (n=21) are presented as scatter blue dot plots. Both groups were tested at 2- and 8-months post two doses of BNT162b2 vaccination (mpv) indicated on the graph as Vac2. Additionally, 15/21 of COVID-19 convalescent vaccinated and 12/21 of COVID-19 naïve vaccinated were tested at 1.5 months post Moderna mRNA-1273 vaccination indicated on the graph as Vac3. Neutralization potency of serum samples were calibrated using WHO International Standard 20/136 and are presented on the graph as International Units per ml [IU/ml]. Solid red lines and numbers correspond to geometric mean in vaccinated COVID-19 convalescent individuals. Solid blue lines and numbers correspond to geometric mean in vaccinated COVID-19 naïve individuals. Dashed line marks the cut-off titer for neutralization assay (45 IU/ml). NT50 was calculated from individual results obtained by triplicates using a nonlinear regression model fit with settings for log inhibitor versus normalized response curves by GraphPad Prism v8. ns: not significant; *p<0.05; **p<0.01; ***p<0.001. Abbreviations: mpv, months postvaccination; International Units per ml [IU/ml]; Vac2, 2 doses of BNT162b2 vaccination; Vac3, third dose of Moderna mRNA-1273 vaccination

**Figure 2.**
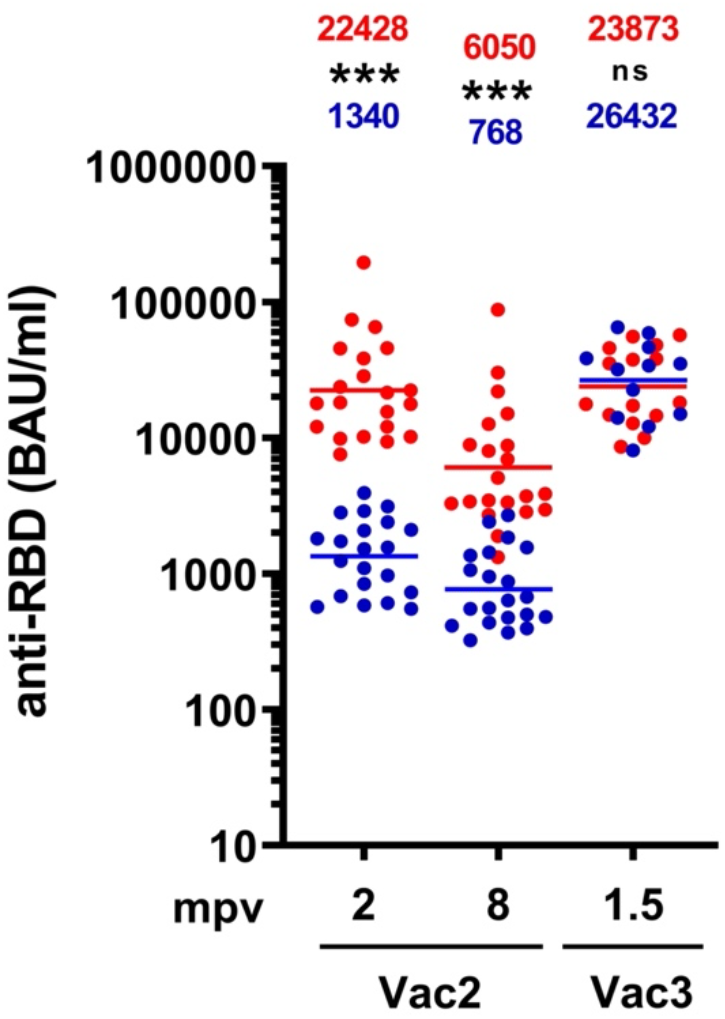
SARS-CoV-2 RBD-binding antibodies in vaccinated COVID-19 convalescent and naïve individuals. COVID-19 convalescent vaccinated (n=21) individuals are presented as scatter red dot plots and COVID-19 naïve vaccinated (n=21) are presented as scatter blue dot plots. Both groups were tested at 2- and 8-months post two doses of BNT162b2 vaccination (mpv) indicated on the graph as Vac2. Additionally, 15/21 of COVID-19 convalescent vaccinated and 12/21 of COVID-19 naïve vaccinated were tested at 1.5 months post Moderna mRNA-1273 vaccination indicated on the graph as Vac3. Solid red lines and numbers correspond to geometric mean in vaccinated COVID-19 convalescent individuals. Solid blue lines and numbers correspond to geometric mean in vaccinated COVID-19 naïve individuals. RBD-binding antibody titers are presented as BAU/ml. ns: not significant; *p<0.05; **p<0.01; ***p<0.001. Abbreviations: mpv, months postvaccination; IgG, immunoglobulin G; BAU/ml, binding antibody units per ml; Vac2, 2 doses of BNT162b2 vaccination; Vac3, third dose of Moderna mRNA-1273 vaccination

## Discussion

A third dose of an mRNA vaccine resulted as expected in an increase of the neutralizing response whose potency and breadth were more significant in naïve individuals as compared with COVID-19 convalescent patients. This later group, as described(7), experienced a great neutralizing response with a wide coverage against SARS-CoV-2 VoC including Omicron after the first two doses. A third dose of an mRNA vaccine restored neutralizing levels in convalescents after a waning process developed during the subsequent months. The effect of a third dose in naïve individuals induced a significant increase of neutralizing titers against the reference SARS-CoV-2 sequence that exceeded the levels achieved after the first two doses (5-fold, p<0.0001) and in a similar range of titers achieved by convalescent (GMT 2469 and 3148 IU/mL respectively). Neutralizing titers against VoC Alpha, Beta, Gamma and Delta exhibited the same pattern of a significant increase in the naïve group as compared with convalescent (p<0.001). In a similar way, the neutralizing activity against Omicron, from a very low titer with the first two doses after the third dose was GMT 832 IU/ml, also in the same range as the level of convalescents: GMT 894 IU/mL. These results support the fact that repeated events of immune stimulation either by natural infection and/or vaccination are required to achieved maximum levels of neutralizing response with a wide breadth against SARS-CoV-2. The number of stimulation events is likely limited to three, since a third dose of an mRNA vaccine did not significantly increase the neutralizing levels achieved after two doses in COVID-19 convalescent patients, as it has been recently observed(8). In the naïve group the third dose of mRNA vaccine induced a clear boosting of neutralizing response in terms of potency and breadth that reached that was comparable for the first time to the levels of convalescents vaccinated. Affinity maturation of antibodies against SARS-CoV-2 Spike protein induced by repeated exposures to infection by pre-VoC SARS-CoV-2 and/or mRNA vaccines based in the ancestral sequence appears to play a role in the increased breadth of neutralization demonstrated in both groups against Omicron(9). However, maturation affinity of antibodies after natural infection and vaccination, in the so-called hybrid immunity(10), appears to be superior. This is highlighted by the fact the neutralizing levels against the related but distant SARS-CoV-1 were significantly higher in the convalescent group as compared with naïves (GMT 768 vs 397 IU/mL respectively, p=0.018). COVID-19 vaccines and specially those based in mRNA confer durable protection against severe disease and death upon SARS-CoV-2 infection. This is most probably based in the stimulation of cell-mediated immunity that seems to be well preserved against the circulating VoC(11). However, at the upper respiratory tract mucosal level, neutralizing antibodies are the main response that is able to block infection and the level of neutralizing antibodies is considered the best correlate of protection for COVID-19 vaccines(12). This response can be outsmarted by the great capability of SARS-CoV-2 to accommodate changes in critical neutralizing epitopes within the spike protein and particularly the RBD region. Three doses of mRNA COVID-19 vaccines, being the third stimulus the most potent, achieved high levels of neutralizing antibodies with a reasonable coverage of VoC including the highly mutated Omicron. Based in the results obtained in convalescent patients undergoing the same vaccination schedule it does not appear that subsequent vaccine doses will achieve further potency. Affinity maturation of antibodies after repeated stimuli, even induced by the ancestral SARS-CoV-2 sequence, allows a wide coverage against VoC. This is especially significant in convalescent-vaccinated that exhibit high levels of neutralization against the related SARS-CoV-1. These results have implication in the development of long-term vaccination strategies.

## Data Availability

All data produced in the present study are available upon reasonable request to the authors

## Conflict of Interest Statement

All authors declare not having conflicts of interests related to this work

## Funding Statement

This work was supported by by grants from the Instituto de Investigación Carlos III, ISCIII, (FIS PI1801007 and PI2100989), by the European Commission Horizon 2020 Framework Programme: Project VIRUSCAN FETPROACT-2016: 731868, Horizon Europe Framework programme: Project EPIC-CROWN-2 ID: 101046084 and by Fundación Caixa-Health Research (Project StopEbola HR18-00469) to RD. M.M-L holds a clinical research contract Río Hortega (CM19/00226) from the Instituto de Salud Carlos III, ISCIII (Spanish Ministry of Science, Innovation and Universities).

## Acknowledgments

We are greatly thankful to all participants in the Solidarity II cohort study

## Notes

### Competing Interest Statement

The authors have declared no competing interest.

### Funding Statement

This work was supported by by grants from the Instituto de Investigacin Carlos III, ISCIII, (FIS PI1801007 and PI2100989), by the European Commission Horizon 2020 Framework Programme: Project VIRUSCAN FETPROACT-2016: 731868, Horizon Europe Framework programme: Project EPIC-CROWN-2 ID: 101046084 and by Fundacion Caixa-Health Research (Project StopEbola HR18-00469) to RD. M.M-L holds a clinical research contract Rio Hortega (CM19/00226) from the Instituto de Salud Carlos III, ISCIII (Spanish Ministry of Science, Innovation and Universities)

### Author Declarations

Comite Etico de Investigacion Medica (IRB) Hospital Universitario 12 de Octubre. Ref CEIm 20/157

